# Anti-Sars-Cov-2 IgA And IgG In Human Milk After Vaccination Is Dependent On Vaccine Type And Previous Sars-Cov-2 Exposure: A Longitudinal Study

**DOI:** 10.1101/2021.05.20.21257512

**Authors:** Marta Selma-Royo, Christine Bäuerl, Desirée Mena-Tudela, Laia Aguilar-Camprubí, Francisco J Pérez-Cano, Anna Parra-Llorca, Carles Lerin, Cecilia Martínez-Costa, Maria Carmen Collado, on behalf of MilkCORONA study team

**Affiliations:** Department of Biotechnology, Institute of Agrochemistry and Food Technology-National Research Council (IATA-CSIC), Paterna, Valencia, Spain; Department of Nursing, Nursing Research Group, Universitat Jaume I, Castellón, Spain; LactApp Women Health, Barcelona, Spain; Physiology Section, Department of Biochemistry and Physiology, Faculty of Pharmacy and Food Science and Institute of Research in Nutrition and Food Safety (INSA), University of Barcelona (UB), Barcelona, Spain; Health Research Institute La Fe, Neonatal Research Group, Spain and University and Polytechnic Hospital La Fe, Division of Neonatology, Valencia, Spain; Institut de Recerca Sant Joan de Déu, Hospital Sant Joan de Déu, Barcelona, Spain; Department of Pediatrics, Hospital Clínico Universitario, University of Valencia, Spain. Nutrition Research Group of INCLIVA, Valencia, Spain

**Keywords:** breast milk, SARS-CoV-2, antibodies, immunoglobulins, vaccines

## Abstract

**Importance:** Limited data are available on COVID-19 vaccine impact in lactating women.

**Objective:** To evaluate the impact of different COVID-19 vaccines on specific anti-SARS-CoV-2 IgA and IgG levels in human milk.

**Design, Settings and Participants:** In this prospective observational study in Spain, 75 lactating women from priority groups receiving vaccination against SARS-CoV-2 were included (January to April 2021). Human milk samples were collected at seven-time points. A group with confirmed SARS-CoV-2 infection (n=19) and a group of women from prepandemic time (n=13) were included.

**Exposure:** mRNA vaccines (BNT162b2 and mRNA-1273) and adenovirus-vectored vaccine (ChAdOx1 nCoV-19).

**Main Outcome(s) and Measure(s):** Presence of IgA and IgG against RBD SARS-CoV-2 in breast milk.

**Results:** Seventy-five vaccinated lactating women [mean age, 34.9 ± 3.7 years] of whom 96% were Caucasic and 92% were health care workers. A total of 417 milk samples were included and vaccine distribution was BioNTech/Pfizer (BNT162b2, n=30), Moderna (mRNA-1273, n=21), and AstraZeneca (ChAdOx1 nCoV-19, n=24). For each vaccine, 7 time points were collected from baseline up to 25 days after the 1^st^ dose and same points were collected for mRNA vaccines 30 days after 2^nd^ dose. A strong reactivity was observed for IgG and IgA after vaccination mainly after the 2^nd^ dose. Presence and the persistence of specific SARS-CoV-2 antibodies in breast milk were dependent on the vaccine-type and, on previous virus exposure. High inter-variability was observed, being relevant for IgA antibodies. IgG levels were significantly higher than those observed in milk from COVID-19 women while IgA levels were lower. Women with previous COVID-19 increased the IgG levels after the 1^st^ dose to a similar level observed in vaccinated women after the 2^nd^ dose.

**Conclusions and Relevance:** Breast milk from vaccinated women contains anti-SARS-CoV-2 IgA and IgG, with highest after the 2nd dose. Levels were dependent on vaccine type and previous exposure to SARS-CoV-2. Previous COVID-19 influenced the vaccine effect after a single dose, which could be especially relevant in the design of vaccination protocols. Further studies are warranted to demonstrate the potential protective role of these antibodies against COVID-19 in infants from vaccinated and infected mothers through breastfeeding.

**Trial Registration:** NCT04751734

**Key Points:** *Question:* What is the effect of the different COVID-19 vaccines on the anti-SARS-CoV-2 antibodies in breast milk? Is the vaccine-specific antibody response in milk comparable to a natural infection? What would be the effect of vaccination on human milk antibodies in women with past SARS-CoV-2 infection?

*Findings:* In this prospective, observational and multicenter study in Spain, lactating women within the priority groups receiving the vaccination against SARS-CoV-2 were included. Although there is a high intra- and inter-variability in the generation of specific SARS-CoV-2 antibodies in breast milk, they are also dependent on the vaccine-type and previous viral exposure.

*Meaning:* Maternal SARS-CoV-2 vaccination provides anti-SARS-CoV-2 antibodies, both IgA and IgG, in human milk and it depends on vaccines and previous COVID-19.

## Introduction

Breastfeeding is the most important postnatal link between mothers and infants, being the best source of nutrition with effects for infant health and development^1^ including the maturation of the neonatal immune system, which is especially relevant in the context of Coronavirus Disease 2019 (COVID-19) vaccination. Similarly to other infective processes^2^ some studies have reported the presence of specific antibodies in milk after SARS-CoV-2 infection. In particular, a rapid and strong antibody response is induced after maternal SARS-CoV-2 infection, with the subsequent accumulation of substantial amounts of specific neutralizing secretory IgA (sIgA) and other antibody types in breast milk ^3–5^.

Europe initiated the vaccination program against COVID-19 on December 27^th^, 2020. In Spain, health care workers were priority groups to receive the vaccines, including breastfeeding women. First available vaccines were mRNA-based vaccines BNT162b2 mRNA and mRNA-1273; later on, an adenovirus-vectored vaccine (ChAdOx1 nCoV-19) became available. While these vaccines protect against severe COVID-19 disease in adult populations ^6–8^, limited data are available on lactating women as they were not included in vaccine trials^1^. Despite this lack of information, main organizations including the Centers for Disease Control and Prevention have recommended lactating women to be immunized ^9,10^. Preliminary studies showed that mRNA-based vaccines induced anti-SARS-CoV-2 antibodies in breast milk ^10–12^. However, several questions remain still open including the extend of the vaccination effect, whether there is a differential response depending on the vaccine type, and the impact of vaccination on women with past SARS-CoV-2 infection.

We investigated whether maternal immunization with the available vaccines resulted in secretion of antibodies directed against SARS-CoV-2 into breast milk and evaluated any potential adverse events among women and their infants. Furthermore, we compared milk antibody levels from vaccinated women to those found in naturally immunized women.

## Methods

### Study population and design

This is a prospective observational, longitudinal, and multicenter study in lactating women receiving vaccination against SARS-CoV-2 infection in Spain (ClinicalTrials.gov Identifier: NCT04751734). Recruitment period started in January 2021 and it is still ongoing. Participants were breastfeeding women within the vaccination priority groups (frontline health care workers) established by the Spanish Government. Participants were recruited at hospitals and health care centers as well as by using specific tools as LactApp^13^ and social media. Women were excluded if cessation of breastfeeding and/or vaccine side-effects required specific treatment and/or hospitalization. Participants received information and written consent was obtained before enrollment. All procedures were in accordance with the ethical standards approved by the Ethical Committee of the Hospital Clínico Universitario (Ref. 2020/133), the Institut de Recerca Sant Joan de Déu (Ref. PIC-94-21), and CSIC (Ref. 061/2021).

Participants received either two doses of mRNA vaccines (BNT162b2 mRNA, BioNTech/Pfizer and mRNA-1273, Moderna) or just one dose of adenovirus-vectored vaccine (ChAdOx1 nCoV-19, Oxford/AstraZeneca). Human milk samples were collected longitudinally at seven-time points: pre-vaccination (1-T0: 0 weeks, 1 week (1-T1), 2 weeks (1-T2) and 3 weeks (1-T3) post the 1^st^ dose of vaccine; and 1 week (2-T1), 2 weeks (2-T2) and 3-4 weeks (2-T3) post 2^nd^ dose for mRNA vaccines. For the adenovirus-vectored vaccine, samples of the 1^st^ vaccine dose follow-up were collected. The vaccination time of the 2^nd^ dose of this last vaccine is still uncertain due to side effects issues, it is being evaluated for other target-population groups and is not provided systematically to all people vaccinated with a first dose.

Furthermore, a group with confirmed SARS-CoV-2 infection (n=19) (ClinicalTrials.gov Identifier: NCT04768244) from a previous study^14^ and a control group of women not exposed to SARS-CoV-2 from prepandemic time^15^ (n=13) (ClinicalTrials.gov Identifier: NCT03552939) were included.

### Human milk collection and processing

Breast milk collection was performed by each women at home following the recommended procedures ^16^. Then, milk was collected mainly by use of a sterile pumper in sterile bottles to normalize the collection among participants and morning collection was recommendable. Breast milk samples were stored immediately at −20 °C and later stored at −80 °C as detailed elsewhere^14^. In brief, samples were thawed and centrifuged, the whey milk was aliquoted and frozen at −80 °C until use. COVID19 positive and prepandemic milk samples were processed in same manner.

### Detection of specific SARS-CoV-2-antibodies in breast milk

Antibodies against the receptor-binding domain (RBD) SARS-CoV-2 S protein were determined as previously described^14^. 1:4 diluted samples were incubated for 2 h in 1 % (w/v) milk powder and SARS-Cov-2 antibodies were detected using anti-human IgA (α-chain-specific) HRP antibody (Thermo-Fisher Scientific; A18781; 1:6.000) and anti-human IgG (Fc specific) HRP antibody (Sigma-Aldrich; A0170; 1:4.000). Standards for interpolation of arbitrary units (AU) were created by pooling 10 breast milk samples with high OD values based on preliminary data. 10 samples from the final collection time (2-T3) of vaccinated mothers and COVID-19 positive mothers were selected for IgG and IgA, respectively. An arbitrary antibody concentration unit of 3000 was assigned to the highest OD value corresponding to the undiluted sample. The standard curve corresponds to eight 3-fold serial dilution from 3000 to 1.38 arbitrary units. In the case of IgG levels, an additional standard curve using a monoclonal anti-SARS-CoV-2 IgG (Clone CR3022 produced in *Nicotiana benthamiana*, BEI Resources, NR-53876) was obtained to assess the equivalence between the arbitrary units and IgG quantification (**eFigure 1**).

**Figure 1.**
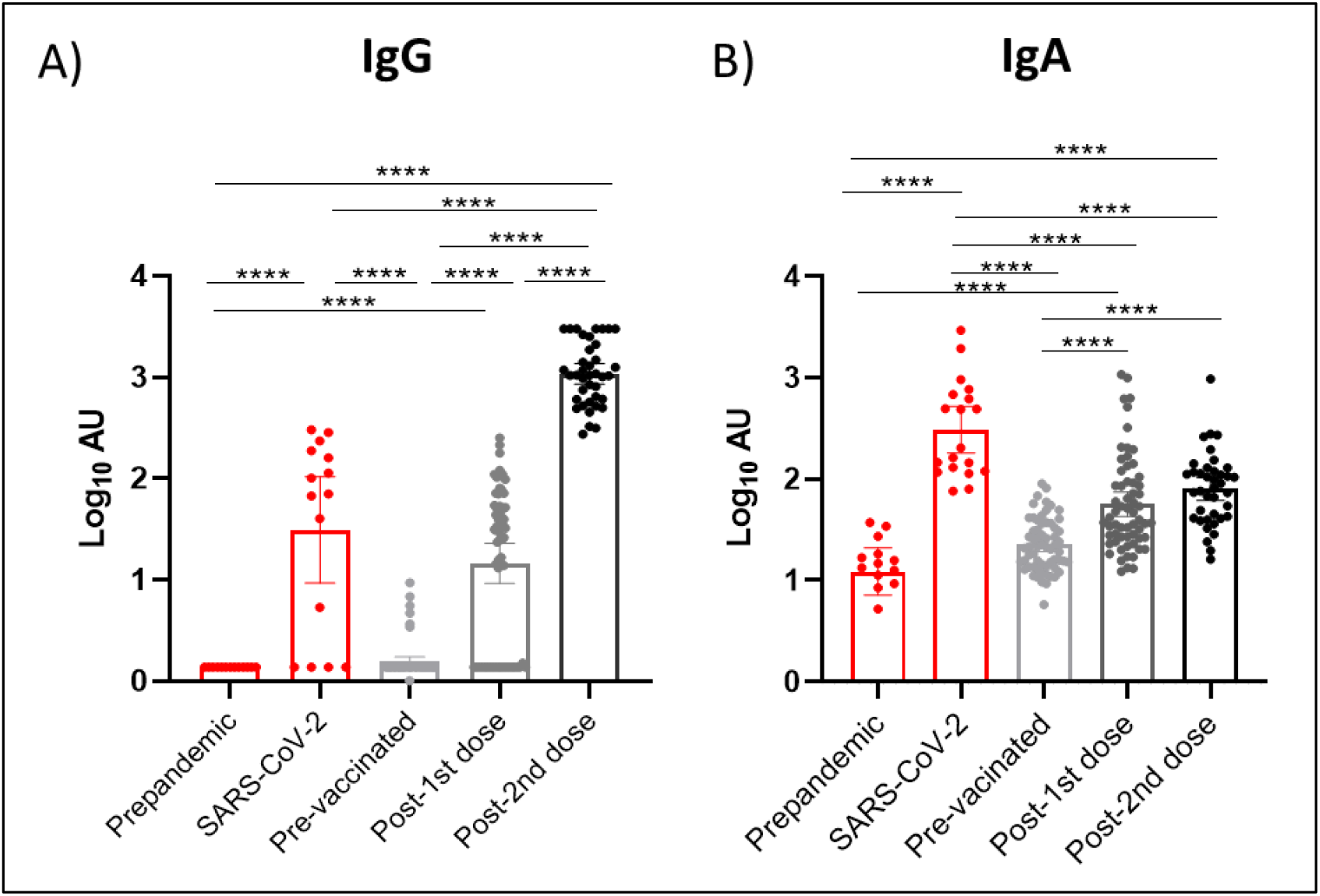
Effect of vaccination on IgG and IgA presence in breast milk. Results of IgG (A) and IgA (B) levels of the three available vaccines in Spain were grouped according to the post-vaccination days. Time points shown in the figure are as following: Post-1^st^ dose (2 weeks after 1^st^ dose of the three studied vaccines), post-2^nd^ dose (2 week after 2^nd^ dose of the mRNA-based vaccines). One-way ANOVA with a Tukey’s post-hoc test for multiple comparisons was performed to assess the statistical significance between groups. Data is presented as mean and 95% of CI of the log-transformed arbitrary units (AU). * p<0.05, ** p<0.01, *** p<0.001, **** p<0.0001.

### Statistical analysis

GraphPad Prism 8.4.3 was used for the statistical analysis. Antibody kinetics were fitted using a nonlinear 4-parameter least-square fit in GraphPad Prism 8.4.3. The resulted fit was used to model the tested samples calculating the arbitrary units of each sample. The maximum (3000 AU) or lowest (1.37 AU) values were assigned to those samples that could not be modelized because of they were out of range in the OD values in the upper or the bottom part of the standard curve, respectively. To compare differences in AU, data were log transformed. Mixed-effects analyses to accommodate missing values were performed to model longitudinal trends of anti-SARS-CoV-2 IgG and IgA. Extended description of the statistical methods is in Supplementary data.

## Results

### Study population characteristics

The study included seventy-five lactating women receiving COVID-19 vaccination (**Table 1**), with age of 34.9±3.9 years (mean±SD) and infant age of 13.4±7.3 months (mean±SD). No significant differences were observed for the vaccine-type groups in clinical variables (**Table 1**). Mothers vaccinated with the adenoviral-vectored vaccine reported more side effects after vaccination compared to those reported with the mRNA-based vaccines including fever (p=0.0005) and headache (p<0.0001) (**eTable 1**). Four infants developed fever after maternal vaccination. There were no serious adverse events during the study period.

**Table 1:** Characteristics of the volunteers included in the study.

|  | BioNtech/Pfizer<br>N=30 | Moderna<br>N=21 | Oxford/AstraZeneca<br>N=24 | p-value |
| --- | --- | --- | --- | --- |
| **Maternal characteristics** |  |  |  |  |
| Age (y) | 34.2 ±4.3 | 35.3 ±3.7 | 34.6 ±2.4 | 0.838 |
| Chronic diseases<br>(hypothyroidism, heart<br>disease) | 6 (20.0%) | 4 (19.0%) | 3(12.5%) | 0.583 |
| Medication | 5 (16.7%) | 3 (14.3%) | 2 (8.3%) | 0.817 |
| Gestational age (weeks) | 39.1 ±2.3 | 39.6 ±1.4 | 39.9 ±1.3 | 0.259 |
| Education degree (health-<br>workers) | 6 (20.0%) | 20 (95.2%) | 20 (83.3%) | <0.0001* |
| SARS-CoV-2 positive PCR<br>before vaccine | 6 (20.0%) | 2 (9.5%) | 0 (0%) | 0.066 |
| **Infant characteristics** |  |  |  |  |
| Gender (female, %) | 9 (30.0%) | 9 (42.8%) | 12 (50.0%) | 0.313 |
| Mode of birth (C-section, %) | 6 (20.0%) | 6 (28.6%) | 3 (12.5%) | 0.440 |
| Weight at birth (g) | 3295.5 ±416.0 | 3500.3 ±600.5 | 3445.2 ±371.5 | 0.260 |
| Length at birth (cm) | 50.8 ±2.1 | 50.6 ±2.4 | 51.2 ±1.8 | 0.617 |
| Infant age at maternal<br>vaccination (month) | 13.2±8.4 | 11.4±5.5 | 14.7±7.0 | 0.319 |
| Exclusive breastfeeding at<br>maternal vaccination (%) | 3 (10.0%) | 3 (14.3%) | 1 (4.2%) | 0.501 |

Categorical data is presented as positive cases (% of the total population). Chi-squared test and T-test were used to test significant differences in clinical categorical and continuous variables, respectively, according to vaccine.

### Anti-SARS-CoV-2 reactive antibodies in breast milk after vaccination

A strong reactivity was observed for IgG and IgA after vaccination mainly after the 2^nd^ dose (**Figure 1**). Both IgG (**Figure 1A**) and IgA (**Figure 1B**) reached higher levels than those in the prepandemic group and those in baseline time-point. Mixed-effects analysis also revealed that vaccination increased IgG (p<0.0001) and IgA (p<0.0001) levels in milk samples (**eTable S2**). While no differences were observed at baseline compared to prepandemic samples, 8 participants who reported SARS-CoV-2 infection prior to vaccination showed higher antibody levels compared to the baseline levels. Four other participants showed the same antibody profile but without confirmation of previous infection. These 4 participants were not included in further analyses.

IgG levels after the 2^nd^ dose of the mRNA-based vaccines reached higher levels than those observed in the SARS-CoV-2 group (p<0.0001) (**Figure 1A**), suggesting a strong response to the vaccines to generate IgG antibodies. Regarding IgA, low levels of non-specific binding were observed in both prepandemic and baseline milk samples (**Figure 1B**). While anti-SARS-CoV-2 IgA levels significantly increased following the 1^st^ and the 2^nd^ dose (p<0.0001 and p<0.0001, respectively), they stayed lower than those found in the SARS-CoV-2 group (p<0.0001).

### Anti-SARS-CoV-2 antibody levels in milk according to vaccine type

We next compared the immunogenic response for each vaccine type. While all three vaccines increased anti-SARS-CoV-2 IgG (**Figure 2A**) and IgA levels (**Figure 2B**), BioNtech/Pfizer and Moderna induced higher IgG levels than Oxford/AstraZeneca after administration of the 1^st^ dose. mRNA-based vaccines induced their maximum effect 2 weeks after 2^nd^ dose (**Figure 2A and 2B, e Figure 2**). Furthermore, a higher percentage of samples from Moderna and BioNtech/Pfizer remained positive based on the stablished cut-off compared to Oxford/AstraZeneca 2 weeks after the 1^st^ dose, (p<0.0001) (**eTable 3**). No major differences were found between the two mRNA-based vaccines, and all samples were classified as positive after the 2^nd^ dose (**eTable 3**).

**Figure 2.**
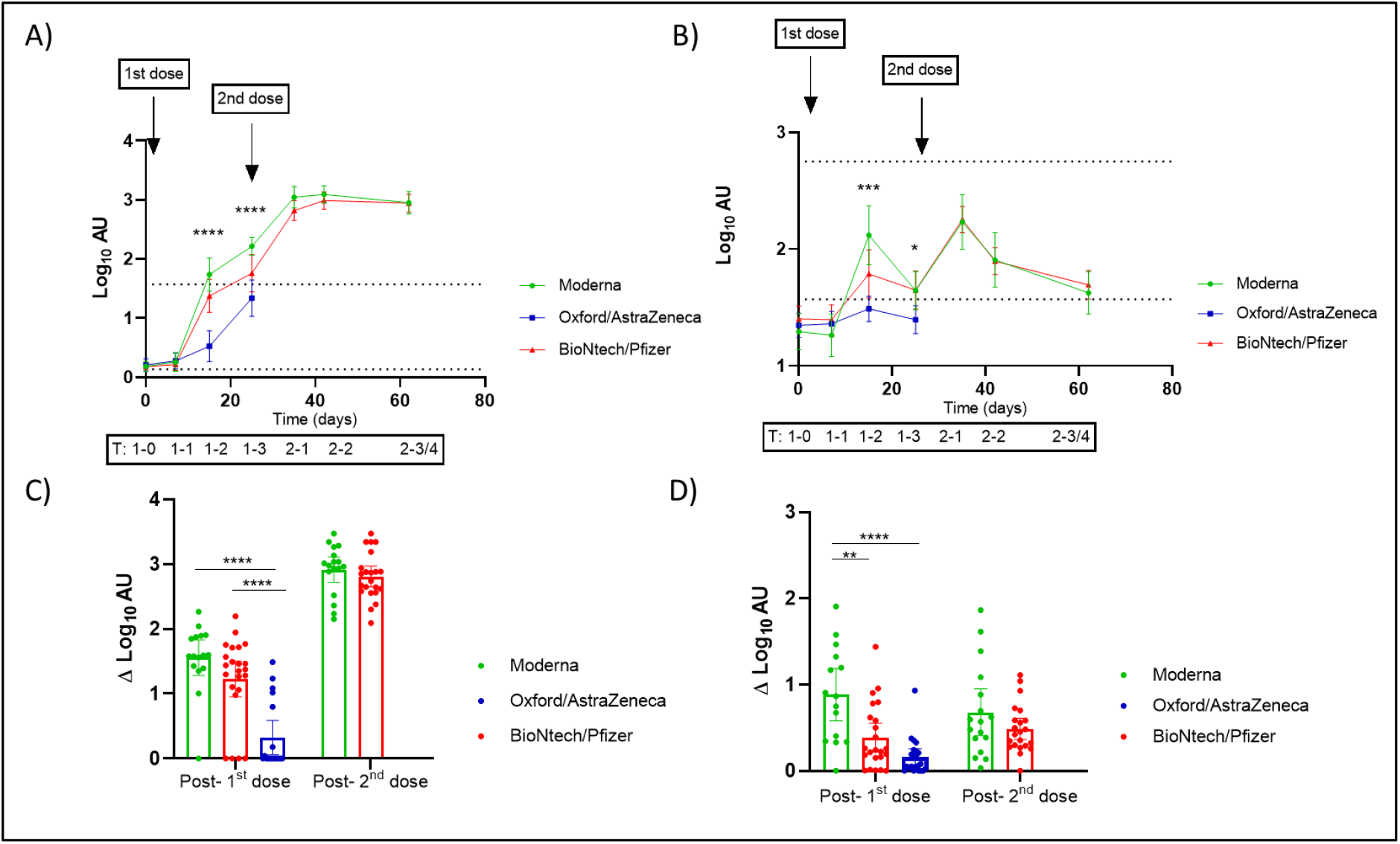
Effect of vaccination on IgG and IgA found in breast milk according to the administered vaccines in Spain. **A, B)** Trajectories of the anti-SARS-CoV-2 IgG (A) and IgA (B) in breast milk samples according to vaccine from baseline (before the 1^st^ dose) to 3-4 weeks post vaccination course. For the adenovirus-vectored vaccine (Oxford/AstraZeneca) only the period up to 3 weeks following the 1^st^ dose was analyzed. The upper dotted line in the plots of trajectories represent the mean of the log transformed arbitrary units (AU) for mothers confirmed of SARS-CoV-2 infection. The lower dotted lines in the trajectories plot represent the established positive cut-off value (log of the mean+2SD of prepandemic anti-SARS-CoV-2 IgA and IgG AU). Mixed-effect analysis with multiple comparisons post-hoc test was performed to test the significance in the antibody’s presence in breast milk and the differences according to vaccines (**Supplementary data, Table S4**). Y-axis marked the approximated days of sampling and the code of the timing as following: pre-vaccination (1-T0: 0 days), 1 week (1-T1), 2 weeks (1-T2) and 3 weeks (1-T3) post the 1st dose of vaccine; and 1 week (2-T1), 2 weeks (2-T2) and 3-4 weeks (2-T3/4) post 2nd dose. Comparison of the increment in log-transformed AU from the determination of SARS-CoV-2 IgG (C) and IgA (D) from baseline to 2 weeks post 1^st^ dose and 2^nd^ dose. One-way ANOVA with a Tukey’s post-hoc test for multiple comparisons was performed to assess the statistical significance of the differences in the antibody detection. Data is presented as mean and 95% of CI of the log-transformed arbitrary units (AU). * p<0.05, ** p<0.01, *** p<0.001, **** p<0.0001.

Mixed-effects analysis for the 1^st^ dose period revealed an effect of vaccine-type on both IgG and IgA levels (p<0.0001), with higher immunoglobulin levels achieved after vaccination with mRNA-based Moderna and BionTech/Pfizer compared to adenoviral-vectored Ofxord/AstraZeneca vaccines at 2- and 3-weeks post 1^st^ dose (**eTable 4**).

We then calculated the increase in IgG and IgA levels from baseline to 2 weeks after 1^st^ dose (for the three vaccines) and 2 weeks after 2^nd^ dose (for mRNA-based vaccines) (**Figure 2C and 2D)**. After the 1^st^ dose, Moderna and BioNtech/Pfizer vaccinated mothers showed higher increment of milk anti-SARS-CoV-2 IgG compared to Oxford/AstraZeneca (p<0.0001), while anti-SARS-CoV-2 IgA levels were higher in Moderna compared to Oxford/AstraZeneca (p<0.0001) and BioNtech/Pfizer (p=0.002) groups. After the 2^nd^ dose, no differences were observed between the two mRNA-based vaccines (**Figures 2C and 2D**). Moreover, after the 2^nd^ dose, IgG levels reached higher levels compared to the 1^st^ dose while IgA levels did not further increase.

### Effects of vaccination in mothers with previous SARS-CoV-2 infection

The subset of mothers with previous SARS-CoV-2 infection (n=8) showed higher IgG and IgA levels at baseline when compared to the prepandemic group and participants without infection (**Figure 3**). Baseline anti-SARS-CoV-2 IgG levels in milk were similar than those observed in non-vaccinated and infected women (SARS-CoV-2) and higher than prepandemic group. IgG levels increased after the 1^st^ dose reaching values similar to those observed after the 2^nd^ dose in participants without previous exposure to SARS-CoV-2 (**Figure 3A**). Baseline anti-SARS-CoV-2 IgA levels were lower compared to the positive control group (p<0.0001), reaching similar values after the 1^st^ dose of the vaccine (**Figure 3B**).

**Figure 3.**
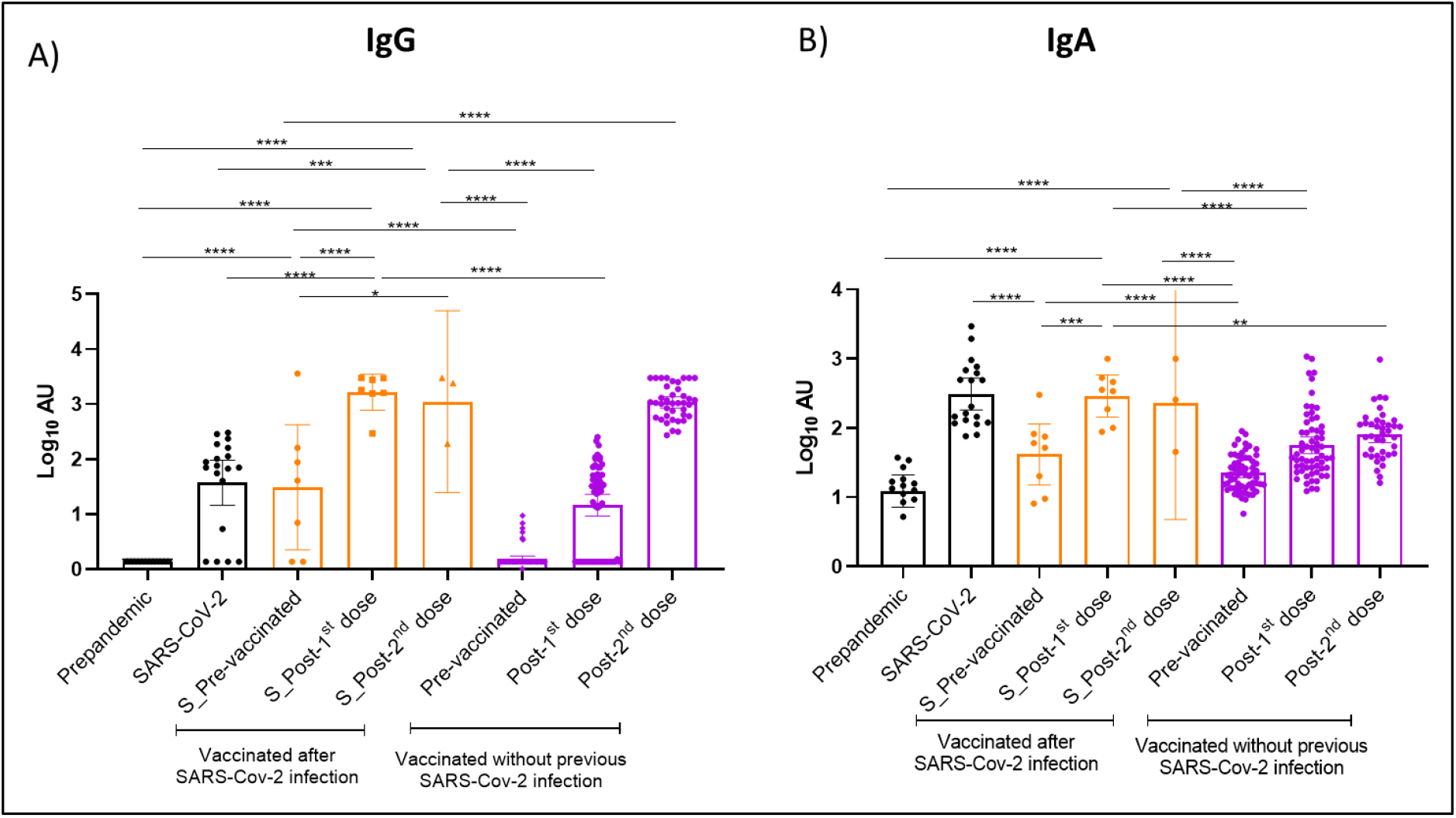
Effect of SARS-CoV-2 vaccination on antibody presence in breast milk in mothers with previous SARS-CoV-2 infection compared to those without previous exposure. Maternal samples from SARS-CoV-2 positive women (orange) before vaccination and those without viral exposure before vaccination (purple) were analyzed for detection of IgG (A) and IgA (B) antibody levels against RBD SARS-CoV-2. One-way ANOVA with a Tukey’s post-hoc test for multiple comparisons was performed to assess the statistical significance of the differences in the antibody detection. Only those differences between the grouped samples of the vaccinated mothers with previous SARS-CoV-2 infection and all other analyzed groups are shown. The time points shown in the figure are as following: Post-1^st^ dose (2 weeks after 1^st^ dose of the three studied vaccines), post-2^nd^ dose (2 weeks after 2^nd^ dose of the mRNA-based vaccines). Data is presented as mean and 95% of CI of the log-transformed arbitrary units (AU). * p<0.05, ** p<0.01, *** p<0.001, **** p<0.0001.

## Discussion

This study reports presence of anti-SARS-CoV-2 IgA and IgG in breast milk after vaccination, determined they were filling the gap on the knowledge about a potential vaccine-dependent immunogenic response. This study represents the biggest study up to now in Europe reporting specific antibody levels in milk for three different vaccines.

It has been reported that COVID-19 mRNA vaccination in pregnant and lactating women generated a robust serum immune response ^10^. However, limited data are available on the impact of the different COVID-19 vaccines in breast milk. Breast milk acts as the main route of passive immunity from mother to infant after birth, which could be especially relevant in the pandemic context. Even though breast milk is considered the best nutrition source for infant development during the first months of live, there were uncertain guidelines for lactating women at the beginning of the pandemic. Presence of IgG and other specific antibodies against SARS-CoV-2 including IgA have been well studied after COVID-19 disease in serum and breast milk^17,18^. However, information on their dynamics after vaccination remains limited. While accumulating data are being provided for mRNA-based vaccines ^12,19,20^, no information is available for the adenovirus-based vaccine (AstraZeneca).

Our study shows increased levels of both anti-SARS-VoV-2 IgG and IgA levels after vaccination. The immunogenic response showed high intra- and inter-individual variability and was dependent on the vaccine type. Both IgA and IgG significantly increased at 2-weeks after the 1^st^ dose in all the analyzed vaccines and showed another peak after the 2^nd^ dose. While levels of anti-SARS-CoV-2 IgG were maintained at the final time point of the follow-up, IgA levels decreased 3-4 weeks after the 2^nd^ dose. These results are in accordance with previous studies focused on the presence of SARS-CoV-2 antibodies in breast milk ^11,20^. Indeed, we found that all samples remained positive for anti-SARS-CoV-2 IgG at the end of the vaccination course, while only 50-60% of the samples were classified as positive for IgA, which is in agreement with previously reported data^12^. Thus, our observations support vaccination as a useful clinical strategy to affect anti-SARS-CoV-2 IgG levels also in breast milk with the potential protective effect for the infant. Further studies with longer follow-up periods are needed for a deeper description of vaccination effects on antibody detection in breast milk as well as for the potential preventive effects in infants and to confirm the clinical relevance of the findings. We observed a robust secretion of SARS-CoV-2 specific IgGs in breast milk after the 2^nd^ dose of vaccination, reaching higher levels than in naturally SARS-CoV-2 infected women, suggesting a powerful effect of vaccination on SARS-CoV-2 IgG production. In agreement with our data, a 2^nd^ mRNA vaccine dose increased the levels of anti-SARS-CoV-2 IgG but not IgA in both maternal blood and breast milk ^10^.

IgA secretion was evident as early as 2 weeks after vaccination followed by a peak after 4 weeks (a week after the 2^nd^ vaccine). Afterwards, levels of specific IgA against RBD gradually decrease during the 3-4 weeks post-vaccination. However, while anti-SARS-CoV-2 IgG levels were higher than those found in the SARS-CoV-2 group, IgA was lower compared to naturally infected women. Further studies with longer follow-up are needed to track persistence of IgA specific antibodies. Indeed, we reported higher baseline signal for anti-SARS-CoV-2 IgA than those found for IgG, which agrees with previous studies ^14^. In this sense, a recent study reported higher S1 + S2-reactive IgA in breast milk from women that had viral respiratory infection symptoms before the pandemic than in milk from women without symptoms ^21^. These data would partially explain the potential signal or cross-reactivity in breast milk samples before 2020 ^21^.

While different studies have reported presence of specific IgA antibodies against SARS-CoV-2 in human milk after COVID-19 ^3–5,17^, to our knowledge none of the previous studies included the different vaccine-types in the same analysis. While we observed no significant differences between the two mRNA-based vaccines, anti-SARS-CoV-2 IgA and IgG levels in women receiving an adenoviral-vectored vaccine were lower compared to mRNA-based vaccines at least after the 1^st^ dose. However, these results need to be carefully considered since our study included data from samples collected up to the 1^st^ dose of Oxford/AstraZeneca vaccine which could not provide a complete view of the vaccination effect in terms of antibody detection in breast milk. Indeed, we could not conclude the clinical relevance of this difference since it is well known that not only antibody but also other immune components including antigen specific T-cells could be activated by vaccines and play a role in the protection conferred by the adenoviral-vectored vaccines ^22^. Nonetheless, our findings highlight that further studies with bigger sample size and independent populations are needed in order to confirm our observations. We also described the effect of vaccination in a small subset of mothers with a previous SARS-CoV-2 infection. Even though the small sample size, these mothers showed a high baseline IgG signal, similar to the positive control group and to that reported after the 1^st^ dose in mothers with no previous infection. Indeed, mothers with a previous SARS-CoV-2 infection reached the peak of anti-SARS-CoV-2 IgG levels after the 1^st^ dose, and these were comparable to those reached in the general studied population after the 2^nd^ dose of the vaccination course. Our results suggest that mothers with previous infection could get a similar potential protective effect in their milk after a unique dose of the vaccination course. Recently, similar results in serum samples from health care workers have been reported ^23^, showing that specific IgG antibodies directed to different SARS-CoV-2 antigens (S1, S2, RBD, and N regions) persisted 3 weeks after a single vaccination^23^. Other studies, in agreement with our results, demonstrated that a single dose of mRNA-based vaccines elicited rapid immune responses in seropositive participants with previous history of SARS-CoV-2 exposure ^24,25^. However, the persistence and duration of antibody responses in both milk and blood need further investigation. Nonetheless, further studies with larger sample size focused on mothers with an history of SARS-CoV-2 infection could be crucial to confirm our results and to clarify the clinical relevance of these findings as well as the potential consequence for the vaccination programs.

## Limitations

Our study provides preliminary data, and it has limitations. Larger prospective studies, bigger samples sizes and distinct cohorts in different locations are needed to confirm the safety of SARS-CoV-2 vaccination in breastfeeding women and the possible differences in the presence of antibodies in breast milk according to vaccine type. Our study does not provide data after the 2^nd^ dose of the adenoviral-vectored vaccine which would be especially relevant in the comparison among the vaccine types. In addition, longer follow-up of the antibody levels would be needed in order to determine their persistence. Furthermore, although it has been suggested that antibodies found in breast milk would exert strong neutralizing effects, no functional assays were performed. In addition, the potential impact on neonatal growth as well as the potential protective effect against infection in the infant remains elusive.

## Conclusions

Breast milk from vaccinated women contains specific IgA and IgG against SARS-CoV-2 RBD, with levels increasing considerably after a 2^nd^ dose in the case of IgG. Women with previous COVID-19 history increased antibody levels y after the 1^st^ dose to the level observed in vaccinated women with no previous infection after the 2^nd^ dose. Further studies are needed to demonstrate the protective antibody effect against COVID-19 in infants from vaccinated and/or infected mothers and the differences in the vaccine-type.

## Data Availability

-

## Acknowledgements

We thank all the families who were involved in the study during this difficult time and in the middle of the COVID-19 pandemic as well as the collaborators of the MilkCORONA (detailed below), which includes neonatologists, pediatricians, midwifes, nurses, research scientists, and computer/laboratory technicians.

MilkCORONA Collaborators:

i) Erika Cortés-Macias (MsC) and Julian Beltrán (Technician) from Department of Biotechnology, Institute of Agrochemistry and Food Technology-National Research Council (IATA-CSIC),Valencia, Spain.

ii) Laura Martínez-Rodríguez (Pediatrician, MD, PhD), Javier Estañ-Capell (Pediatrician, MD, PhD) from Pediatric Nutrition Research Group of INCLIVA Biomedical Research Institute of Valencia and Department of Pediatrics, Hospital Clínico Universitario, University of Valencia, Spain.

iii) Álvaro Solaz-García (RN), Inmaculada Lara-Cantón (MD), Health Research Institute La Fe, Neonatal Research Group, Spain and University and Polytechnic Hospital La Fe, Division of Neonatology, Valencia, Spain.

iv) Victoria Fumadó (MD, PhD), Cristina Garcia (MsC), Cristina Plou (MsC), Institut de Recerca Sant Joan de Déu, Hospital Sant Joan de Déu, 08950 Barcelona, Spain.

v) Maria José Rodríguez-Lagunas (PhD), Karla Río-Aigé (MsC), Physiology Section, Department of Biochemistry and Physiology, Faculty of Pharmacy and Food Science, University of Barcelona (UB), 08028 Barcelona, Spain.

vi) Alba Padró-Arocas (IBCLC) and Paola Quifer-Rada (PhD). LactApp Women Health, Barcelona, Spain.

## Contributions

Concept and design: Martínez-Costa and Collado

Clinical aspects including recruitment of the participants, collection of samples and clinical records: Mena-Tudela, Aguilar-Camprubí, Parra-Llorca, Lerin, Martínez-Costa. Acquisition, analysis, or interpretation of data: Selma-Royo, Bäuerl, Pérez-Cano, Lerin, Martínez-Costa, Collado

Drafting of the manuscript: Selma-Royo, Bäuerl, Collado

Critical revision of the manuscript for important intellectual content: Selma-Royo, Bäuerl, Mena-Tudela, Aguilar-Camprubí, Pérez-Cano, Parra-Llorca, Lerin, Martínez-Costa, Collado

## Conflict of Interest Disclosures

None reported.

## Notes

### Competing Interest Statement

The authors have declared no competing interest.

### Clinical Trial

NCT04751734

### Funding Statement

-

### Author Declarations

All procedures were in accordance with the ethical standards approved by the Ethical Committee of the Hospital Clinico Universitario (Ref. 2020/133), the Institut de Recerca Sant Joan de Deu (Ref. PIC-94-21) and CSIC (Ref. 061/2021).

## References

1. Mena-Tudela D, Aguilar-Camprubí L, Quifer-Rada P, Paricio-Talayero JM, Padró-ArocasThe COVID-19 vaccine in women: Decisions, data and gender gap. Nurs Inq. Published online March 28, 2021:e12416. doi:10.1111/nin.12416

2. Schlaudecker EP, Steinhoff MC, Omer SB, et al. IgA and neutralizing antibodies to influenza a virus in human milk: a randomized trial of antenatal influenza immunization. PLoS One. 2013;8(8):e70867. doi:10.1371/journal.pone.0070867

3. Fox A, Marino J, Amanat F, et al. The Spike-specific IgA in milk commonly-elicited after SARS-Cov-2 infection is concurrent with a robust secretory antibody response, exhibits neutralization potency strongly correlated with IgA binding, and is highly durable over time. medRxiv. Published online March 20, 2021:2021.03.16.21253731. doi:10.1101/2021.03.16.21253731

4. Pace RM, Williams JE, Järvinen KM, et al. Characterization of SARS-CoV-2 RNA, Antibodies, and Neutralizing Capacity in Milk Produced by Women with COVID-19. mBio. 2021;12(1). doi:10.1128/mBio.03192-20

5. Favara DM, Ceron-Gutierrez ML, Carnell GW, Heeney JL, Corrie P, Doffinger R. Detection of breastmilk antibodies targeting SARS-CoV-2 nucleocapsid, spike and receptor-binding-domain antigens. Emerg Microbes Infect. 2020;9(1):2728–2731. doi:10.1080/22221751.2020.1858699

6. Baden LR, El Sahly HM, Essink B, et al. Efficacy and Safety of the mRNA-1273 SARS-CoV-2 Vaccine. N Engl J Med. 2021;384(5):403–416. doi:10.1056/NEJMoa2035389

7. Polack FP, Thomas SJ, Kitchin N, et al. Safety and Efficacy of the BNT162b2 mRNA Covid-19 Vaccine. N Engl J Med. 2020;383(27):2603–2615. doi:10.1056/NEJMoa2034577

8. Ramasamy MN, Minassian AM, Ewer KJ, et al. Safety and immunogenicity of ChAdOx1 nCoV-19 vaccine administered in a prime-boost regimen in young and old adults (COV002): a single-blind, randomised, controlled, phase 2/3 trial. Lancet. 2021;396(10267):1979–1993. doi:10.1016/S0140-6736(20)32466-1

9. CDC. Vaccination Considerations for People Pregnant or Breastfeeding. Centers for Disease Control and Prevention. Published April 28, 2021. Accessed May 9, 2021. https://www.cdc.gov/coronavirus/2019-ncov/vaccines/recommendations/pregnancy.html

10. Gray KJ, Bordt EA, Atyeo C, et al. COVID-19 vaccine response in pregnant and lactating women: a cohort study. Am J Obstet Gynecol. Published online March 24, 2021. doi:10.1016/j.ajog.2021.03.023

11. Perl SH, Uzan-Yulzari A, Klainer H, et al. SARS-CoV-2-Specific Antibodies in Breast Milk After COVID-19 Vaccination of Breastfeeding Women. JAMA. Published online April 12, 2021. doi:10.1001/jama.2021.5782

12. Fox A, Norris C, Amanat F, Zolla-Pazner S, Powell RL. The vaccine-elicited immunoglobulin profile in milk after COVID-19 mRNA-based vaccination is IgG-dominant and lacks secretory antibodies. medRxiv. Published online January 1, 2021:2021.03.22.21253831. doi:10.1101/2021.03.22.21253831

13. Padró-Arocas A, Quifer-Rada P, Aguilar-Camprubí L, Mena-Tudela D. Description of an mHealth tool for breastfeeding support: LactApp. Analysis of how lactating mothers seek support at critical breastfeeding points and according to their infant’s age. Res Nurs Health. 2021;44(1):173–186. doi:10.1002/nur.22095

14. Bäuerl C, Randazzo W, Sánchez G, et al. SARS-CoV-2 RNA and antibody detection in human milk from a prospective multicenter study in Spain. medRxiv. Published online January 1, 2021:2021.05.06.21256766. doi:10.1101/2021.05.06.21256766

15. García-Mantrana I, Alcántara C, Selma-Royo M, et al. MAMI: a birth cohort focused on maternal-infant microbiota during early life. BMC Pediatr. 2019;19(1):140. doi:10.1186/s12887-019-1502-y

16. McGuire MK, Seppo A, Goga A, et al. Best Practices for Human Milk Collection for COVID-19 Research. Breastfeed Med. 2021;16(1):29–38. doi:10.1089/bfm.2020.0296

17. Demers-Mathieu V, Do DM, Mathijssen GB, et al. Difference in levels of SARS-CoV-2 S1 and S2 subunits-and nucleocapsid protein-reactive SIgM/IgM, IgG and SIgA/IgA antibodies in human milk. Journal of Perinatology. 2021;41(4):850–859. doi:10.1038/s41372-020-00805-w

18. Keulen BJ van, Romijn M, Bondt A, et al. Breastmilk; a source of SARS-CoV-2 specific IgA antibodies. medRxiv. Published online August 21, 2020:2020.08.18.20176743. doi:10.1101/2020.08.18.20176743

19. Esteve-Palau E, Gonzalez-Cuevas A, Guerrero ME, et al. Quantification of specific antibodies against SARS-CoV-2 in breast milk of lactating women vaccinated with an mRNA vaccine. medRxiv. Published online April 7, 2021:2021.04.05.21254819. doi:10.1101/2021.04.05.21254819

20. Valcarce V, Stafford LS, Neu J, et al. Detection of SARS-CoV-2 specific IgA in the human milk of COVID-19 vaccinated, lactating health care workers. medRxiv. Published online January 1, 2021:2021.04.02.21254642. doi:10.1101/2021.04.02.21254642

21. Demers-Mathieu V, DaPra C, Mathijssen G, et al. Human Milk Antibodies Against S1 and S2 Subunits from SARS-CoV-2, HCoV-OC43, and HCoV-229E in Mothers with A Confirmed COVID-19 PCR, Viral SYMPTOMS, and Unexposed Mothers. Int J Mol Sci. 2021;22(4). doi:10.3390/ijms22041749

22. Ewer KJ, Barrett JR, Belij-Rammerstorfer S, et al. T cell and antibody responses induced by a single dose of ChAdOx1 nCoV-19 (AZD1222) vaccine in a phase 1/2 clinical trial. Nat Med. 2021;27(2):270–278. doi:10.1038/s41591-020-01194-5

23. Bradley T, Grundberg E, Selvarangan R, et al. Antibody Responses after a Single Dose of SARS-CoV-2 mRNA Vaccine. N Engl J Med. Published online March 23, 2021. doi:10.1056/NEJMc2102051

24. Krammer F, Srivastava K, Alshammary H, et al. Antibody Responses in Seropositive Persons after a Single Dose of SARS-CoV-2 mRNA Vaccine. N Engl J Med. 2021;384(14):1372–1374. doi:10.1056/NEJMc2101667

25. Levi R, Azzolini E, Pozzi C, et al. One dose of SARS-CoV-2 vaccine exponentially increases antibodies in recovered individuals with symptomatic COVID-19. J Clin Invest. Published online May 6, 2021. doi:10.1172/JCI149154

