## Supplementary material for "Anti-Sars-Cov-2 IgA And IgG In Human Milk After Vaccination Is Dependent On Vaccine Type And Previous Sars-Cov-2 Exposure: A Longitudinal Study": Suplementary material

**SUPPLEMENTARY DATA**

**Extended statistical methods**

For the study of the general study of IgA and IgG levels (**Figure 1**), Dunnet’s test for multiple comparison was used as post-hoc test in the mixed-effect analysis to compare IgA and IgG levels at 15 days after 1st and 2nd doses with the baseline condition. For the analysis of the percentage of positive samples, milk samples were classified as positive or negative considering the positive cut-off values for each Ig class calculated from the AU of the prepandemic control samples and defined as mean plus two standard deviations (SD). Chi-square test and Fisher’s exact test was used to analyze differences in the percentage of positive samples according to vaccines.

For trajectory studies, two mixed-effects analysis were independently performed to model 1) from baseline to 21-25 days post 1st dose with the three vaccines (Tukey post-hoc test); and 2) the complete course of vaccination for the two mRNA-based vaccines (Sidak post-hoc test). Other statistical evaluations were assessed using one-way ANOVA with Tukey’s post-hoc test.

Maternal, pregnancy, and birth characteristics were collected as covariates for descriptive purposes and matched on maternal characteristics for associations between SARS-CoV-2 positive women and prepandemic groups, and neonatal outcomes. Chi-square test was used to assess potential associations between vaccine type and side-effects. A p-value<0.05 was considered statistically significant.

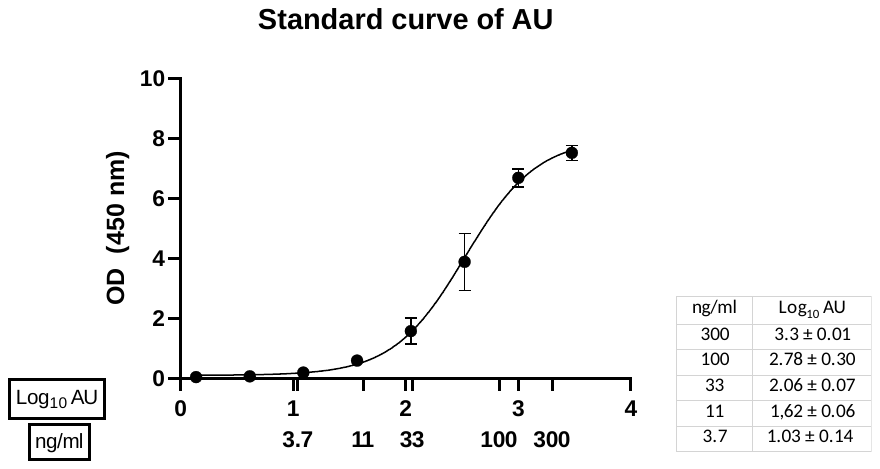

**eFigure 1.** Equivalence of the standard curve composed of pooled milk samples to the monoclonal anti-SARS-CoV-2 IgG, clone CR3022. The table shows the mean ± SD of the log-transformed arbitrary units that correspond to each concentration of monoclonal anti-SARS-CoV-2 IgG.

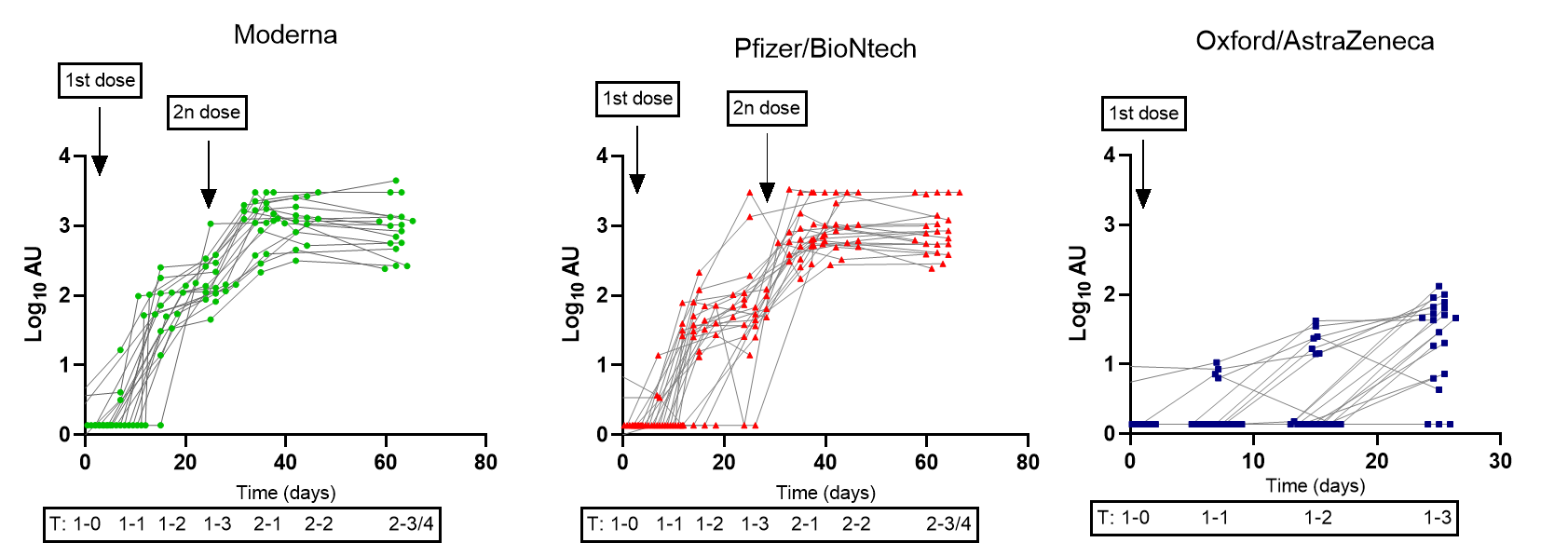

**eFigure 2.** Individual trajectories of the SARS-CoV-2 IgG in breast milk samples according to vaccine from baseline (before the 1^st^ dose) to 3-4 weeks post vaccination course. For the adenovirus-vectored vaccine (Oxford/AstraZeneca) only the first 3 weeks following the 1^st^ dose were analyzed. Data is presented as log-transformed arbitrary units (AU).

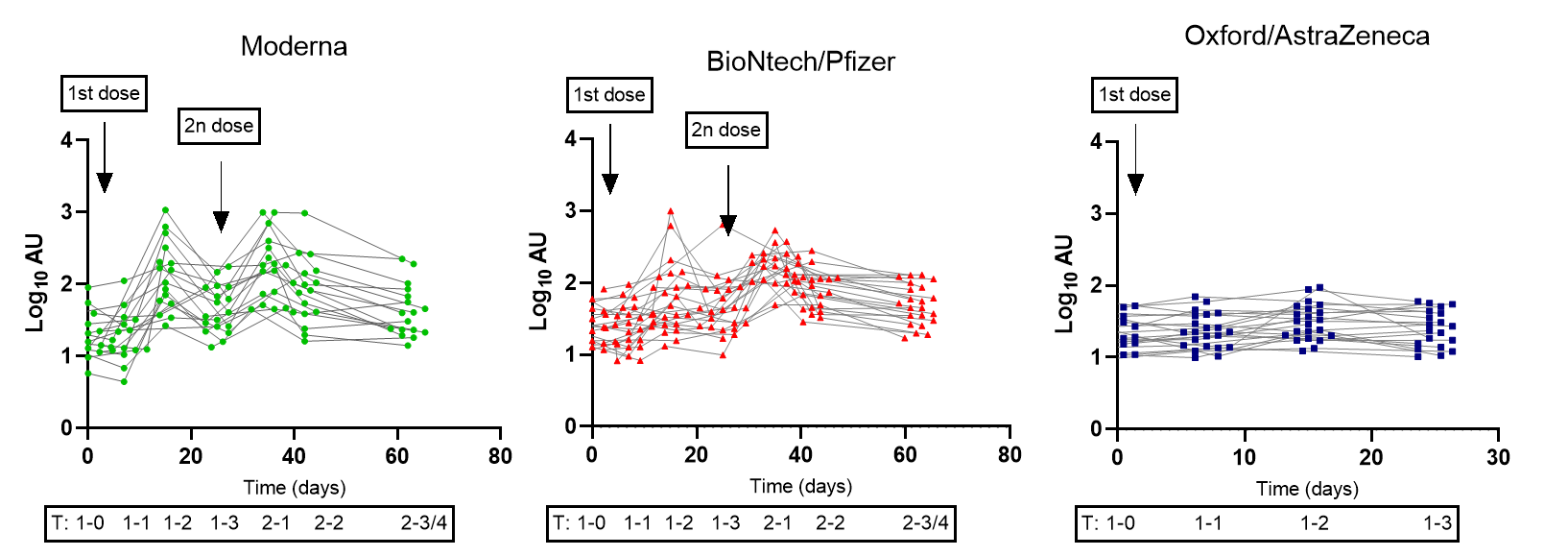

**eFigure 3.** Individual trajectories of the SARS-CoV-2 IgA in breast milk samples according to vaccine from baseline (before the 1^st^ dose) to 3-4 weeks post vaccination course. For the adenovirus-vectored vaccine (Oxford/AstraZeneca) only the first 3 weeks following the 1^st^ dose were analyzed. Data is presented as log-transformed arbitrary units (AU).

**eTable 1.** Women self-reported side-effects after vaccination and infant effects.

|  | BioNtech/Pfizer  N=30 | Moderna  N=21 | Oxford/AstraZeneca  N=24 | p-value |
| --- | --- | --- | --- | --- |
| Maternal side-effects | | | | |
| 1st dose |  |  |  |  |
| Local pain and tiredness | 27 (90.0%) | 19 (90.5%) | 23 (95.8%) | 0.702 |
| Fever | 2 (6.7%) | 0 (0.0%) | 9 (37.5%) | **0.0005*** |
| Headache | 4 (13.3%) | 5 (23.8%) | 18 (75.0%) | **<0.0001*** |
| Others (myalgia, insomnia, nausea) | 2 (6.7%) | 4 (19.0%) | 11 (45.8%) | **0.003*** |
| 2nd dose |  |  |  |  |
| Local pain and tiredness | 28 (93.3%) | 20 (95.2%) | - | >0.999 |
| Fever | 10 (33.3%) | 6 (28.6%) | - | 0.768 |
| Headache | 14 (46.7%) | 6 (28.6%) | - | 0.250 |
| Others (myalgia, insomnia, nausea) | 11 (36.6%) | 9 (42.8%) | - | 0.773 |
| Infant side-effects |  |  |  |  |
| 1st dose |  |  |  |  |
| Skin reaction (dermatitis, etc) | 0 (0.0%) | 1 (4.8%) | 2 (8.3%) | 0.293 |
| Fever | 1 (3.3%) | 0 (0.0%) | 0 (0.0%) | 0.468 |
| Irritability and insomnia | 2 (6.7%) | 0 (0.0%) | 6 (25.0%) | **0.017*** |
| 2nd dose |  |  |  |  |
| Skin reaction | 0 (0.0%) | 0 (0.0%) | - | - |
| Fever | 3 (10.0%) | 2 (9.5%) | - | >0.999 |
| Irritability and insomnia | 5 (16.7%) | 5 (23.8%) | - | 0.722 |

Data is expressed as positive cases (% of the total population). Chi-square test (three vaccines) and Exact-Fisher’s test (mRNA-based vaccines) were used to the comparisons for the side-effects between the analyzed vaccines.

**eTable 2.** Results from the longitudinal mixed-effects analysis modeling the changes in IgG and IgA detection in human breast milk after vaccination.

|  | Pre-vaccination | Post 1st dose | Post 2nd dose | Baseline vs 1st dose | p-value | Baseline vs  2^nd^ dose | p-value |
| --- | --- | --- | --- | --- | --- | --- | --- |
| SARS CoV-2 IgA | 1.35 ± 0.26 | 1.78 ±0.47 | 1.90 ± 0.35 | -0.413  -0.56, -0.27 | <0.0001 | -0.549  -0.68, -0.41 | <0.0001* |
| SARS CoV-2 IgG | 0.19 ± 0.18 | 1.16 ±0.77 | 3.03 ±0.31 | -0.973  -1.20, -0.75 | <0.0001 | -2.84  -2.97, -2.72 | <0.0001* |

Data is presented as log-transformed arbitrary units (AU). The following time points were considered in the model: Post-1st dose (2 weeks after 1st dose of the three studied vaccines), post- 2nd dose (3-4 weeks after 2nd dose of the mRNA-based vaccines). The first three columns present the mean of log-transformed arbitrary units ± SD and the comparisons columns present the mean difference (95% confidence interval).

|  | Moderna  (n=18)ⱡ | BioNtech  Pfizer  (n=23)ⱡ | Oxford  AstraZeneca  (n=22)ⱡ | p-value |
| --- | --- | --- | --- | --- |
| IgG |  |  |  |  |
| 2 weeks  Post 1st dose | 93.8% ^¥^ | 83.3% | 36.4% | **>0.0001*** |
| 3 weeks  Post 1st dose | 100% | 93.3% | 85.0% ^¥^ | 0.205 |
| 2 weeks  Post 2nd dose | 100%^#^ | 100% | - | >0.999 |
| 3-4 weeks  Post 2nd dose | 100%^#^ | 100% | - | >0.999 |
| IgA |  |  |  |  |
| 2 weeks  Post 1st dose | 87.5% ^¥^ | 56.6% | 36.4% | **0.0004*** |
| 3 weeks  Post 1st dose | 50%^#^ | 56.5% | 30% ^¥^ | 0.202 |
| 2 weeks  Post 2nd dose | 82.4%^#^ | 86.4% | - | >0.999 |
| 3-4 weeks  Post 2nd dose | 52.9%^#^ | 63.6% | - | 0.531 |

**eTable 3.** Percentage of mothers with a signal above the stablished cut-off for a positive result for SARS-CoV-2 antibody presence according to vaccine.

ⱡSamples with previous SARS-CoV-2 infection (n=8) and those with a similar antibody profile were removed from the analysis (n=4). The positive cut-off was stablished as a signal above the mean + 2 SD of the arbitrary units from the prepandemic group. Symbols mark the missing values (^#^: One and ^¥^: Two) at the specific time point.

|  | Moderna | BioNtech  /Pfizer | Oxford  AstraZeneca | Moderna vs BioNtech/Pfizer | Moderna vs Oxford | Oxford vs Pfizer |
| --- | --- | --- | --- | --- | --- | --- |
| SARS CoV-2 IgG | | | | | | |
| T1-0 | 0.186 ± 0.17 | 0.179 ± 0.17 | 0.209 ± 0.23 | -0.007  (-0.124, 0.138)  0.991 | -0.023  (-0.181, 0.135)  0.932 | 0.030  (-0.2, 0.180)  0.876 |
| T1-1 | 0.257 ± 0.29 | 0.217 ± 0.23 | 0.276 ± 0.31 | 0.040  (-0.178, 0.259)  0.892 | -0.019  (-0.260, 0.222)  0.979 | 0.060  (-0.138, 0.258)  0.746 |
| T1-2 | 1.738 ± 0.53 | 1.376 ± 0.64 | 0.525 ± 0.59 | 0.361  (-0.097, 0.820)  0.146 | 1.212  (0.768, 1.657)  **<0.0001** | -0.851  (-1.295, -0.407)  **<0.0001** |
| T1-3 | 2.215 ± 0.31 | 1.758 ± 0.72 | 1.337 ± 0.66 | 0.457  (0.045, 0.868)  0.123 | 0.878  (0.473, 1.283)  **<0.0001** | -0.422  (-0.932, 0.089)  **0.027** |
| T2-1 | 3.05 ± 0.35 | 2.828 ± 0.36 |  | 0.228  (-0.107, 0.563)  0.355 |  |  |
| T2-2 | 3.09 ± 0.28 | 2.989 ± 0.33 |  | 0.102  (-0.176, 0.381)  0.919 |  |  |
| T2-3 | 2.95 ± 0.37 | 2.946 ± 0.35 |  | 0.004  (-0.328, 0.337)  >0.999 |  |  |
| SARS CoV-2 IgA | | | | | | |
| T1-0 | 1.294 ± 0.30 | 1.245 ± 0.61 | 1.118 ± 0.47 | -0.109  (-0.331, 0.114)  0.461 | -0.053  (-0.272, 0.167)  0.824 | -0.056  (-0.235, 0.123)  0.732 |
| T1-1 | 1.263 ± 0.34 | 1.225 ± 0.614 | 1.137 ± 0.44 | -0.133  (-0.392, 0.126)  0.425 | -0.097  (-0.345, 0.151)  0.597 | -0.036  (-0.229, 0.158)  0.896 |
| T1-2 | 2.120 ± 0.48 | 2.198 ±1.228 | 1.452 ± 0.55 | 0.333  (-0.048, 0.713)  0.097 | 0.631  (0.303, 0.959)  **0.0002*** | -0.299  (-0.575, -0.022)  **0.032*** |
| T1-3 | 1.647 ± 0.329 | 1.793 ± 0.875 | 1.256 ± 0.60 | -0.002  (-0.272, 0.268)  0.999 | 0.250  (0.013, 0.487)  **0.037*** | -0.253  (-0.491, -0.015)  **0.036*** |
| T2-1 | 2.233 ± 0.455 | 3.331 ± 0.749 |  | 0.495  (-0.711, 1.701)  0.842 |  |  |
| T2-2 | 1.908 ± 0.456 | 2.599 ± 0.809 |  | 0.173  (-0.782, 0.129)  0.998 |  |  |
| T2-3 | 1.626 ± 0.357 | 2.039 ± 0.798 |  | -0.007  (-0.970, 0.955)  >0.999 |  |  |

**eTable 4.** Results from the longitudinal mixed-effects analysis modeling the changes in IgG and IgA detection in human milk at baseline and each analyzed time after vaccination according to vaccine.

Data is presented as log-transformed arbitrary units (AU). Results in the comparison columns are shown as mean difference (95% confidence interval) and p-value. Significant p-values are marked in bold. Two mixed-effects models were performed independently, one for the baseline to 3 weeks post- 1^st^ dose and the other one for the complete vaccination course only for mRNA-based vaccines. Time points are expressed as follows: pre-vaccination (1-T0: 0 days), 1 week (1-T1), 2 weeks (1-T2) and 3 weeks (1-T3) post the 1st dose of vaccine; and 1 week (2-T1), 2 weeks (2-T2) and 3-4 weeks (2-T3) post 2nd dose of vaccine.
